# A time-series analysis of anxiety diagnoses and prescriptions from January 2018 to January 2023

**DOI:** 10.1101/2024.01.22.24301620

**Authors:** Brianna M Goodwin Cartwright, Peter D Smits, Patricia J Rodriguez, Samuel Gratzl, Charlotte Baker, Nick Stucky

**Author notes:** **Corresponding Author:** Nick Stucky, M.D., Ph.D., VP Research, Truveta Inc.

## Abstract

This research aimed to estimate whether first-time anxiety diagnoses and medication usage during and after the COVID-19 pandemic differed from diagnosis and medication usage rates prior to the COVID-19 pandemic, within groups stratified by age. We observed a significant increase in first-time anxiety diagnoses and prescriptions for 12–17-year-olds during April 2020-March 2021 compared to January 2018-February 2020. These trends were not sustained in subsequent periods. We also observed an increase in first-time prescriptions for 18–29-year-olds during the same time. The study emphasizes the importance of addressing mental health concerns, particularly among adolescents, both during and beyond the pandemic.

## Introduction

The COVID-19 pandemic had a profound impact on the mental health of individuals worldwide. Layoffs, furloughs, remote work, homeschooling, and general uncertainty contributed to heightened stress levels and increased psychological distress (Salari et al., 2020). Anxiety can affect many aspects of a person’s life and prolonged anxiety has been linked to physical illness (Clarke & Currie, 2009). Previous studies have explored the prevalence of anxiety symptoms and poor mental health outcomes during the pandemic (Centers for Disease Control and Prevention, 2022; Marin et al., 2023; Pasquini & Keeter, 2022; Woolston, 2020). However, there remains a gap in understanding the specific trends in first-time anxiety diagnoses and medication prescribing patterns, particularly among different age groups.

In this study, we examined whether rates of new anxiety diagnoses or first-time anxiety medication prescribing were higher during the COVID-19 pandemic, compared to pre-pandemic rates. Additionally, we sought to identify any significant variations in first-time diagnosis or prescribing trends within age groups. To achieve these objectives, we compared data from before and during the pandemic.

By focusing on first-time anxiety diagnoses and medication prescriptions, we aim to provide a deeper understanding of the evolving mental health landscape during this unprecedented period. Our study highlights vulnerable groups, such as adolescents, who may have experienced unique challenges and mental health concerns during the pandemic (Imran et al., 2020).

## Methods

We used a subset of Truveta Data to investigate first-time anxiety diagnoses and first-time anxiety medications between January 1, 2018, and January 31, 2023. Truveta Data consists of electronic health records provided by member U.S. healthcare systems that were deidentified by expert determination in accordance with the HIPAA Privacy Rule (Althouse et al., 2023; Goodwin Cartwright et al., 2023; Smits et al., 2023). Anxiety diagnoses were defined using the National Library of Medicine’s Value Set Authority Center definition of Generalized Anxiety Disorder (National Committee for Quality Assurance, 2021). Anxiety medications were defined using the Established Pharmacologic Class provided by the FDA for selective serotonin reuptake inhibitors (SSRI) (FDA, 2021). Both the diagnosis and prescriptions definitions can be found in the **Appendix**. These data were accessed on March 24, 2023.

We calculated the 1) rate of first-time diagnosis per patients with outpatient encounters within each month who had not previous had an anxiety diagnosis and 2) the rate of first-time anxiety prescriptions per patients with outpatient encounters within each month who had not previous received an anxiety prescription. The analyses were conducted independently. We used this as a method to estimate the rate of diagnosis or prescription amongst people who are seeking care. We stratified our population by age (12-17, 18-29, 30-44, 45-64, and 65+) and analyzed each age group independently.

We used the Seasonal-Trend decomposition using LOESS (STL) method to adjust for seasonality in the data using the *STL* function in *statsmodels* python package (Seabold & Perktold, 2010). All graphs are seasonally adjusted and are comprised of the trend and residual components of the time series.

We compared four time periods to test for differences 1) pre-pandemic: January 2018 – February 2020, 2) pandemic onset: March 2020, 3) pandemic period 1: April 2020 – March 2021, and 4) pandemic period 2: April 2021 – January 2023. We allowed the initial pandemic period to have a different effect than the other two pandemic time periods because of the expected difference in healthcare-seeking patterns during March 2020 compared with other months. We fit a linear regression model to the seasonally adjusted data and compared the difference in marginal means between the four time periods using *emmeans* R package (Lenth et al., 2019).

## Results

We identified 34,969,938 and 23,837,222 people who were eligible to have an anxiety diagnosis or medication, respectively. Of those, 2,856,083 (8.2%) had a first-time anxiety diagnosis and 1,818,111 (7.6%) had a first-time anxiety medication prescription between January 2018 and January 2023. **Figure 1** shows the first-time anxiety diagnosis and prescription trends for all age groups. Within all age groups, first-time diagnoses exceeded first-time prescriptions. With the brief exception of early 2021, the rate of first-time diagnosis and prescriptions was highest for the 18-29-year-old age group. Before the pandemic, the 30-44-year-old age group had the second highest rates of both diagnosis and prescriptions. However, after March 2020, the 12-17-year-old age group had a consistently higher first-time prescription rate and occasionally a higher first-time diagnosis rate when compared to the 30-44-year-old age group.

**Figure 1:**
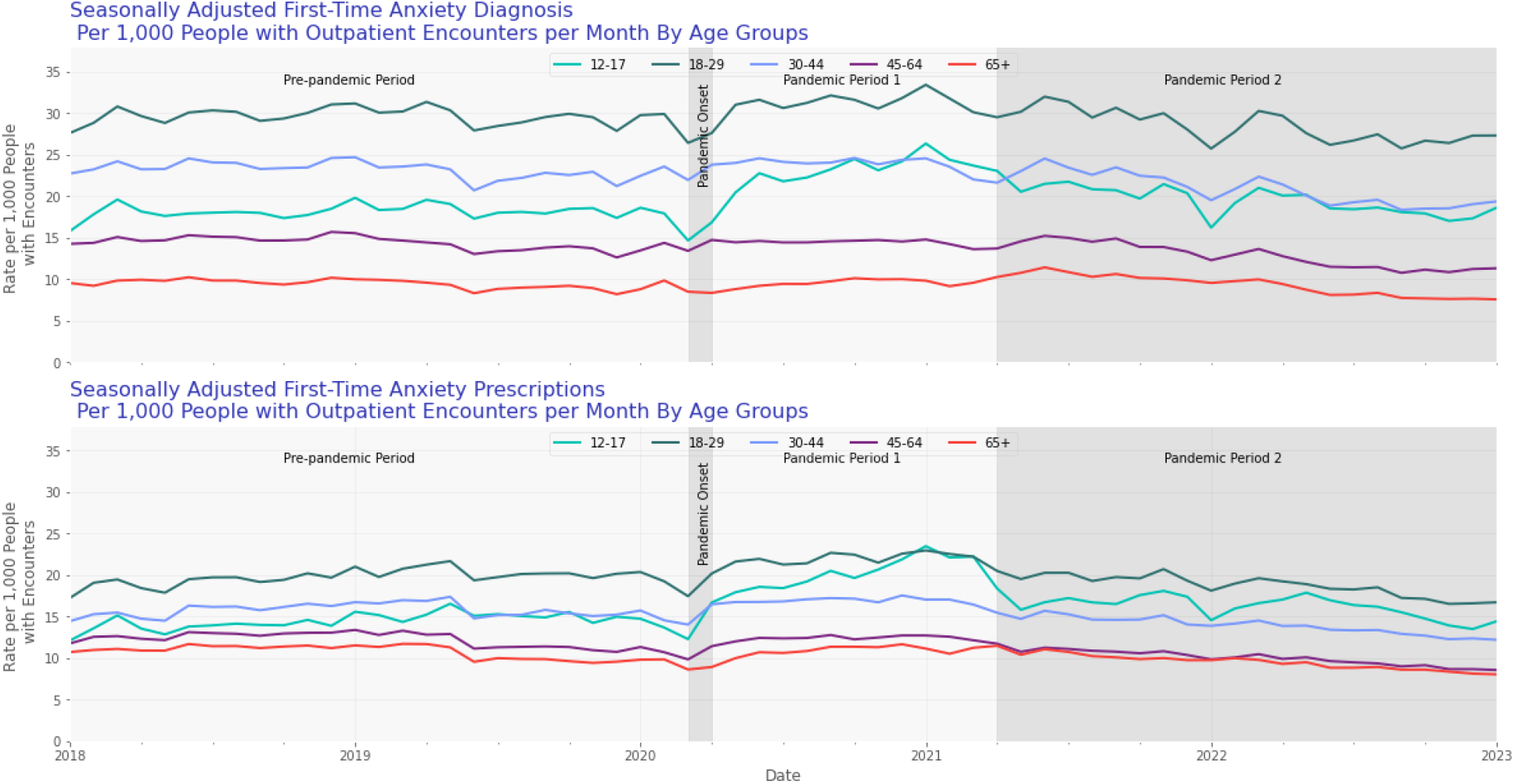
The seasonally adjusted rate of first-time anxiety diagnosis (top) and anxiety medications (bottom) per people with outpatient encounters, without previous diagnosis. The study period is divided into 4 periods: 1) Pre-pandemic period: January 2018 – February 2020, 2) Pandemic Onset: March 2020, 3) Pandemic Period 1: April 2020-March 2021, 4) Pandemic Period 2: April 2021 – January 2023.

For the 12–17-year-old age group we observed a significant increase in first-time anxiety diagnoses (pre-pandemic: 19.0 ± 0.2, pandemic period 1: 21.4 ± 0.4, p<0.001) and anxiety prescriptions (pre-pandemic: 15.5 ± 0.2, pandemic period 1: 18.2 ± 0.4, p<0.001) from the pre-pandemic period to pandemic period 1 (**Table 1**). This trend was not sustained; the pre-pandemic period was not different than pandemic period 2 for this age group (p > 0.05). First-time anxiety prescriptions also increased for 18–29-year-olds, from pre-pandemic compared to both pandemic periods 1 and 2 (p <0.05). First-time diagnoses did not increase during either pandemic period compared with the pre-pandemic period for the 18–29-year-old age group. We did not observe other significant increases in prescription or diagnosis rates when comparing the pandemic periods to the pre-pandemic periods for any group older than 29 years of age.

**Table 1:** The estimated marginal mean rate and standard deviation of first-time anxiety diagnosis and prescriptions by age group and pandemic period.

|  | *12-17 years*<br>*old* | *18-29 years*<br>*old* | *30-44 years*<br>*old* | *45-64 years*<br>*old* | *65+ years*<br>*old* |
| --- | --- | --- | --- | --- | --- |
| **Diagnosis** |  |  |  |  |  |
| *Pre-pandemic*<br>*Period* | $19.0 \pm 0.2$ | $29.6 \pm 0.2$ | $22.7 \pm 0.2$ | $13.9 \pm 0.1$ | $9.4 \pm 0.1$ |
| *Pandemic Onset* | $15.6 \pm 1.1$ | $26.2 \pm 1.1$ | $21.2 \pm 0.9$ | $13 \pm 0.5$ | $8.1 \pm 0.4$ |
| *Pandemic*<br>*Period 1* | $21.4 \pm 0.4$ | $30.3 \pm 0.3$ | $22.8 \pm 0.3$ | $14 \pm 0.2$ | $9.6 \pm 0.1$ |
| *Pandemic*<br>*Period 2* | $19.4 \pm 0.2$ | $28.9 \pm 0.3$ | $22.1 \pm 0.2$ | $13.6 \pm 0.1$ | $9.3 \pm 0.1$ |
| **Medications** |  |  |  |  |  |
| *Pre-pandemic*<br>*Period* | $15.5 \pm 0.2$ | $19.9 \pm 0.2$ | $15.5 \pm 0.1$ | $11.6 \pm 0.1$ | $10.4 \pm 0.1$ |
| *Pandemic Onset* | $13.6 \pm 1$ | $17.8 \pm 0.8$ | $14.5 \pm 0.6$ | $10.4 \pm 0.5$ | $9.0 \pm 0.4$ |
| *Pandemic*<br>*Period 1* | $18.2 \pm 0.4$ | $20.8 \pm 0.3$ | $16.0 \pm 0.2$ | $11.8 \pm 0.1$ | $10.6 \pm 0.1$ |
| *Pandemic*<br>*Period 2* | $16.1 \pm 0.2$ | $19.2 \pm 0.2$ | $14.7 \pm 0.2$ | $11.0 \pm 0.1$ | $10.0 \pm 0.1$ |

## Discussion

The COVID-19 pandemic has had a significant impact on mental health, particularly among vulnerable populations (Amerio et al., 2020; Marin et al., 2023). In this analysis, we estimated the rate of first-time anxiety diagnoses and prescriptions before and during the pandemic. Our findings indicate a noteworthy increase in first-time anxiety diagnoses and medication prescriptions among the 12-17-year-old population during April 2020 – March 2021 (Pandemic Period 1). However, these rates were not sustained in the subsequent period, suggesting the need for continued care and attention for individuals with first-time diagnoses or prescriptions. This research contributes to the existing body of literature by exploring new dimensions of anxiety-related healthcare trends and identifying specific populations at higher risk for impact from effects potentially associated with the pandemic. By better understanding the impact of the pandemic on anxiety diagnoses and medication prescriptions, healthcare professionals and policymakers can develop targeted interventions to support those most in need.

Our findings shed light on the impact of the pandemic on anxiety-related healthcare utilization. Addressing mental health concerns, especially among adolescents, has always been crucial, and the pandemic has further emphasized the importance of prioritizing mental well-being (Choi et al., 2020). Untreated mental health conditions can have long-lasting consequences, impacting various aspects of an individual’s life, including learning abilities, social interactions, and self-esteem (Blakemore, 2019; McCarthy, 2022). Our findings align with previous survey analyses and analyses of inpatient populations outside the United States (Centers for Disease Control and Prevention, 2022; Marin et al., 2023), indicating that adolescents not only reported increased anxiety levels since the onset of the pandemic but also sought and received care at a higher rate.

It is worth noting that our study did not investigate the severity of anxiety symptoms, and further research should explore the potential increase in the severity of anxiety experienced during these challenging periods. Moreover, previous studies have highlighted disparities in anxiety rates among different population groups, based on social determinants of health and lifestyle factors (Centers for Disease Control and Prevention, 2022; Pasquini & Keeter, 2022). Future investigations should consider exploring the impact of these factors, such as essential workers, individuals who had to homeschool their children, or those who experienced furloughs, layoffs, or wage cuts.

Several limitations should be acknowledged in this analysis. Firstly, anxiety and depression are interrelated and share treatment medications, including SSRIs; however, we did not differentiate between the medication used to treat anxiety or depression in this study. Future studies should encompass both anxiety and depression as key outcomes of interest. Additionally, we focused solely on first-time anxiety diagnoses and prescriptions, excluding individuals who may have previously received diagnoses or prescriptions. Therefore, our study likely under captures the full burden anxiety, as it does not include recurrent episodic anxiety or increased severity of prevalent anxiety. Further, people who are taking other medications for anxiety, such as for people who the anxiety medications included are not well tolerated or who may be taking other drugs that interfere with these drugs, were not captured in this study. Furthermore, our dataset consisted of individuals seeking healthcare, thereby potentially excluding those who experienced anxiety but did not engage with the healthcare system.

This analysis contributes to the growing body of research exploring the impact of the COVID-19 pandemic on mental health. By elucidating trends in first-time anxiety diagnoses and medication prescriptions, we identify focused populations that might need additional attention, especially those in the 12-17-year-old age range. Recognizing the evolving landscape of mental health during and after the pandemic, healthcare professionals and policymakers can develop targeted interventions to mitigate the long-term consequences of anxiety and promote holistic well-being.

## Data Availability

The data used in this study are available to all Truveta subscribers and may be accessed at studio.truveta.com.

## Notes

### Competing Interest Statement

All authors were/are employees of Truveta, Inc.

### Funding Statement

This study did not receive any funding.

### Author Declarations

Normalized electronic health record data are de-identified by expert determination under the HIPAA Privacy Rule before being made available to researchers. In accordance with 46.101 Protection of Human Subjects, our study did not require Institutional Review Board approval because it used only deidentified medical records. All data used in this study are publicly available to Truveta subscribers and may be accessed at studio.truveta.com.

