## Appendix for "A time-series analysis of anxiety diagnoses and prescriptions from January 2018 to January 2023"

### Appendix: Anxiety diagnosis and prescription codes used to define first-time anxiety diagnoses or prescriptions.

*Anxiety Diagnosis codes*

| Code System | Concept Code | Concept Name |
|---|---|---|
| **ICD10CM** | F41.0 | Panic disorder [episodic paroxysmal anxiety] |
| **ICD10CM** | F41.1 | Generalized anxiety disorder |
| **ICD10CM** | F41.3 | Other mixed anxiety disorders |
| **ICD10CM** | F41.8 | Other specified anxiety disorders |
| **ICD10CM** | F41.9 | Anxiety disorder, unspecified |
| **SNOMED CT** | 1380006 | Agoraphobia without history of panic disorder with limited symptom attacks |
| **SNOMED CT** | 1686006 | Sedative, hypnotic AND/OR anxiolytic-induced anxiety disorder |
| **SNOMED CT** | 1816003 | Panic disorder with agoraphobia, severe agoraphobic avoidance AND mild panic attacks |
| **SNOMED CT** | 3158007 | Panic disorder with agoraphobia, agoraphobic avoidance in partial remission AND panic attacks in partial remission |
| **SNOMED CT** | 109006 | Anxiety disorder of childhood OR adolescence |
| **SNOMED CT** | 4932002 | Panic disorder with agoraphobia, moderate agoraphobic avoidance AND mild panic attacks |
| **SNOMED CT** | 5509004 | Panic disorder with agoraphobia AND severe panic attacks |
| **SNOMED CT** | 8185002 | Panic disorder with agoraphobia AND moderate panic attacks |
| **SNOMED CT** | 10586006 | Occupation-related stress disorder |
| **SNOMED CT** | 11806006 | Separation anxiety disorder of childhood |
| **SNOMED CT** | 11941006 | Panic disorder with agoraphobia, agoraphobic avoidance in full remission AND panic attacks in full remission |
| **SNOMED CT** | 13438001 | Overanxious disorder of childhood |
| **SNOMED CT** | 17496003 | Organic anxiety disorder |
| **SNOMED CT** | 19766004 | Panic disorder with agoraphobia, mild agoraphobic avoidance AND severe panic attacks |
| **SNOMED CT** | 21897009 | Generalized anxiety disorder |
| **SNOMED CT** | 22230001 | Panic disorder with agoraphobia, agoraphobic avoidance in partial remission AND panic attacks in full remission |
| **SNOMED CT** | 24781009 | Panic disorder with agoraphobia, mild agoraphobic avoidance AND panic attacks in full remission |
| **SNOMED CT** | 25501002 | Social phobia |
| **SNOMED CT** | 30059008 | Panic disorder with agoraphobia, severe agoraphobic avoidance AND moderate panic attacks |
| **SNOMED CT** | 31781004 | Panic disorder with agoraphobia, agoraphobic avoidance in partial remission AND mild panic attacks |
| **SNOMED CT** | 32388005 | Panic disorder with agoraphobia, agoraphobic avoidance in partial remission AND moderate panic attacks |

| | | |
|---|---|---|
| **SNOMED CT** | 34116005 | Panic disorder with agoraphobia, agoraphobic avoidance in full remission AND severe panic attacks |
| **SNOMED CT** | 34938008 | Anxiety disorder caused by alcohol |
| **SNOMED CT** | 35607004 | Panic disorder with agoraphobia |
| **SNOMED CT** | 37868008 | Anxiety disorder of adolescence |
| **SNOMED CT** | 37872007 | Avoidant disorder of childhood OR adolescence |
| **SNOMED CT** | 38328002 | Panic disorder with agoraphobia, severe agoraphobic avoidance AND panic attacks in full remission |
| **SNOMED CT** | 43150009 | Panic disorder without agoraphobia with severe panic attacks |
| **SNOMED CT** | 47505003 | Posttraumatic stress disorder |
| **SNOMED CT** | 49564006 | Panic disorder with agoraphobia, mild agoraphobic avoidance AND moderate panic attacks |
| **SNOMED CT** | 50026000 | Organic anxiety disorder caused by psychoactive substance |
| **SNOMED CT** | 50983008 | Panic disorder with agoraphobia, mild agoraphobic avoidance AND panic attacks in partial remission |
| **SNOMED CT** | 51493001 | Anxiety disorder caused by cocaine |
| **SNOMED CT** | 52910006 | Anxiety disorder due to a general medical condition |
| **SNOMED CT** | 53467004 | Anxiety disorder of childhood |
| **SNOMED CT** | 53956006 | Panic disorder without agoraphobia with panic attacks in partial remission |
| **SNOMED CT** | 54587008 | Simple phobia |
| **SNOMED CT** | 55967005 | Anxiety disorder caused by phencyclidine |
| **SNOMED CT** | 56576003 | Panic disorder without agoraphobia |
| **SNOMED CT** | 58535001 | Physical AND emotional exhaustion state |
| **SNOMED CT** | 59923000 | Panic disorder with agoraphobia AND panic attacks in full remission |
| **SNOMED CT** | 61157009 | Combat fatigue |
| **SNOMED CT** | 61212007 | Panic disorder with agoraphobia, severe agoraphobic avoidance AND severe panic attacks |
| **SNOMED CT** | 61569007 | Agoraphobia without history of panic disorder |
| **SNOMED CT** | 62351001 | Generalized social phobia |
| **SNOMED CT** | 63701002 | Panic disorder with agoraphobia, mild agoraphobic avoidance AND mild panic attacks |
| **SNOMED CT** | 63909006 | Panic disorder with agoraphobia AND panic attacks in partial remission |
| **SNOMED CT** | 64060000 | Panic disorder with agoraphobia, moderate agoraphobic avoidance AND panic attacks in full remission |
| **SNOMED CT** | 64165008 | Avoidant disorder of childhood |
| **SNOMED CT** | 65064003 | Panic disorder without agoraphobia with moderate panic attacks |
| **SNOMED CT** | 67195008 | Acute stress disorder |
| **SNOMED CT** | 69479009 | Anxiety hyperventilation |
| **SNOMED CT** | 70691001 | Agoraphobia |
| **SNOMED CT** | 72861004 | Panic disorder without agoraphobia with mild panic attacks |

| | | |
|---|---|---|
| **SNOMED CT** | 74010007 | Panic disorder with agoraphobia, severe agoraphobic avoidance AND panic attacks in partial remission |
| **SNOMED CT** | 76812003 | Panic disorder with agoraphobia, moderate agoraphobic avoidance AND panic attacks in partial remission |
| **SNOMED CT** | 76868007 | Panic disorder with agoraphobia, agoraphobic avoidance in full remission AND mild panic attacks |
| **SNOMED CT** | 82339009 | Anxiety disorder caused by amfetamine |
| **SNOMED CT** | 82415003 | Agoraphobia without history of panic disorder without limited symptom attacks |
| **SNOMED CT** | 82494000 | Panic disorder without agoraphobia with panic attacks in full remission |
| **SNOMED CT** | 82738004 | Panic disorder with agoraphobia, moderate agoraphobic avoidance AND moderate panic attacks |
| **SNOMED CT** | 83253003 | Shyness disorder of childhood |
| **SNOMED CT** | 83631006 | Panic disorder with agoraphobia, moderate agoraphobic avoidance AND severe panic attacks |
| **SNOMED CT** | 85061001 | Separation anxiety disorder of childhood, early onset |
| **SNOMED CT** | 87798009 | Panic disorder with agoraphobia, agoraphobic avoidance in full remission AND moderate panic attacks |
| **SNOMED CT** | 89948007 | Panic disorder with agoraphobia AND mild panic attacks |
| **SNOMED CT** | 90790003 | Avoidant disorder of adolescence |
| **SNOMED CT** | 111487009 | Dream anxiety disorder |
| **SNOMED CT** | 111490003 | Panic disorder with agoraphobia, agoraphobic avoidance in partial remission AND severe panic attacks |
| **SNOMED CT** | 111491004 | Panic disorder with agoraphobia, agoraphobic avoidance in full remission AND panic attacks in partial remission |
| **SNOMED CT** | 126943008 | Separation anxiety |
| **SNOMED CT** | 191722009 | Agoraphobia with panic attacks |
| **SNOMED CT** | 191724005 | Social phobia, fear of eating in public |
| **SNOMED CT** | 191725006 | Social phobia, fear of public speaking |
| **SNOMED CT** | 191726007 | Social phobia, fear of public washing |
| **SNOMED CT** | 191736004 | Obsessive-compulsive disorder |
| **SNOMED CT** | 191737008 | Compulsive neurosis |
| **SNOMED CT** | 191738003 | Obsessional neurosis |
| **SNOMED CT** | 192014006 | Psychogenic rumination |
| **SNOMED CT** | 192037000 | Acute panic state due to acute stress reaction |
| **SNOMED CT** | 192038005 | Acute fugue state due to acute stress reaction |
| **SNOMED CT** | 192039002 | Acute stupor state due to acute stress reaction |
| **SNOMED CT** | 192041001 | Acute situational disturbance |
| **SNOMED CT** | 192042008 | Acute post-trauma stress state |
| **SNOMED CT** | 192044009 | Stress reaction causing mixed disturbance of emotion and conduct |
| **SNOMED CT** | 192611004 | Childhood phobic anxiety disorder |
| **SNOMED CT** | 197480006 | Anxiety disorder |
| **SNOMED CT** | 231504006 | Mixed anxiety and depressive disorder |

| | | |
|---|---|---|
| **SNOMED CT** | 238965007 | Venereophobia |
| **SNOMED CT** | 238966008 | Syphilophobia |
| **SNOMED CT** | 238976006 | Bromisodrophobia |
| **SNOMED CT** | 279611005 | Shell shock |
| **SNOMED CT** | 313182004 | Chronic post-traumatic stress disorder |
| **SNOMED CT** | 317816007 | Stockholm syndrome |
| **SNOMED CT** | 318784009 | Posttraumatic stress disorder, delayed onset |
| **SNOMED CT** | 371631005 | Panic disorder |
| **SNOMED CT** | 386810004 | Phobic disorder |
| **SNOMED CT** | 403593004 | Phobic fear of skin cancer |
| **SNOMED CT** | 426174008 | Chronic stress disorder |
| **SNOMED CT** | 428687006 | Nightmares associated with chronic post-traumatic stress disorder |
| **SNOMED CT** | 443919007 | Complex posttraumatic stress disorder |
| **SNOMED CT** | 446175003 | Acute posttraumatic stress disorder following military combat |
| **SNOMED CT** | 446180007 | Delayed posttraumatic stress disorder following military combat |
| **SNOMED CT** | 699241002 | Chronic post-traumatic stress disorder following military combat |
| **SNOMED CT** | 723913009 | Olfactory reference disorder |
| **SNOMED CT** | 724654009 | Anxiety disorder caused by opioid |
| **SNOMED CT** | 724693000 | Obsessive compulsive disorder caused by cocaine |
| **SNOMED CT** | 724708007 | Anxiety disorder caused by methylenedioxymethamphetamine |
| **SNOMED CT** | 724722007 | Anxiety disorder caused by dissociative drug |
| **SNOMED CT** | 724723002 | Anxiety disorder caused by ketamine |
| **SNOMED CT** | 724730008 | Obsessive compulsive disorder caused by psychoactive substance |
| **SNOMED CT** | 737341006 | Anxiety disorder caused by synthetic cannabinoid |
| **SNOMED CT** | 762331007 | Anxiety disorder caused by stimulant |
| **SNOMED CT** | 762332000 | Obsessive compulsive disorder caused by stimulant |
| **SNOMED CT** | 762515000 | Anxiety disorder caused by synthetic cathinone |
| **SNOMED CT** | 762516004 | Obsessive compulsive disorder caused by synthetic cathinone |
| **SNOMED CT** | 22621000119103 | Anxiety disorder caused by drug |
| **SNOMED CT** | 10743001000119100 | Anxiety disorder in mother complicating childbirth |
| **SNOMED CT** | 12398201000119100 | Anxiety disorder caused by methamphetamine |
| **SNOMED CT** | 16264621000119100 | Recurrent mild major depressive disorder co-occurrent with anxiety |
| **SNOMED CT** | 16264821000119100 | Recurrent severe major depressive disorder co-occurrent with anxiety |
| **SNOMED CT** | 16264901000119100 | Recurrent moderate major depressive disorder co-occurrent with anxiety |
| **SNOMED CT** | 16265061000119100 | Recurrent major depressive disorder co-occurrent with anxiety in full remission |

| | | |
|---|---|---|
| **SNOMED CT** | 16265301000119100 | Recurrent major depressive disorder in partial remission co-occurrent with anxiety |
| **SNOMED CT** | 16265951000119100 | Mild major depressive disorder co-occurrent with anxiety single episode |
| **SNOMED CT** | 16266831000119100 | Moderate major depressive disorder co-occurrent with anxiety single episode |
| **SNOMED CT** | 16266991000119100 | Severe major depressive disorder co-occurrent with anxiety single episode |

*Anxiety Prescription codes*

| Code System | Concept Code | Concept Name |
|---|---:|---|
| **RxNorm** | 10737 | trazodone |
| **RxNorm** | 2556 | citalopram |
| **RxNorm** | 32937 | paroxetine |
| **RxNorm** | 31565 | nefazodone |
| **RxNorm** | 321988 | escitalopram |
| **RxNorm** | 36437 | sertraline |
| **RxNorm** | 42355 | fluvoxamine |
| **RxNorm** | 4493 | fluoxetine |
| **RxNorm** | 108886 | citalopram 20 MG Oral Tablet [Cipramil] |
| **RxNorm** | 1098648 | nefazodone hydrochloride 100 MG |
| **RxNorm** | 1098649 | nefazodone hydrochloride 100 MG Oral Tablet |
| **RxNorm** | 1098650 | nefazodone hydrochloride 100 MG [Serzone] |
| **RxNorm** | 1098651 | nefazodone hydrochloride 100 MG Oral Tablet [Serzone] |
| **RxNorm** | 1098665 | nefazodone hydrochloride 150 MG |
| **RxNorm** | 1098666 | nefazodone hydrochloride 150 MG Oral Tablet |
| **RxNorm** | 1098667 | nefazodone hydrochloride 150 MG [Serzone] |
| **RxNorm** | 1098668 | nefazodone hydrochloride 150 MG Oral Tablet [Serzone] |
| **RxNorm** | 1098669 | nefazodone hydrochloride 200 MG |
| **RxNorm** | 1098670 | nefazodone hydrochloride 200 MG Oral Tablet |
| **RxNorm** | 1098671 | nefazodone hydrochloride 200 MG [Serzone] |
| **RxNorm** | 1098672 | nefazodone hydrochloride 200 MG Oral Tablet [Serzone] |
| **RxNorm** | 1098673 | nefazodone hydrochloride 250 MG |
| **RxNorm** | 1098674 | nefazodone hydrochloride 250 MG Oral Tablet |
| **RxNorm** | 1098675 | nefazodone hydrochloride 250 MG [Serzone] |
| **RxNorm** | 1098676 | nefazodone hydrochloride 250 MG Oral Tablet [Serzone] |
| **RxNorm** | 1098677 | nefazodone hydrochloride 50 MG |
| **RxNorm** | 1098678 | nefazodone hydrochloride 50 MG Oral Tablet |
| **RxNorm** | 1098679 | nefazodone hydrochloride 50 MG [Serzone] |
| **RxNorm** | 1098680 | nefazodone hydrochloride 50 MG Oral Tablet [Serzone] |

| | | |
|---|---|---|
| **RxNorm** | 1098681 | {14 (nefazodone hydrochloride 100 MG Oral Tablet) / 14 (nefazodone hydrochloride 200 MG Oral Tablet) / 14 (nefazodone hydrochloride 50 MG Oral Tablet) } Pack |
| **RxNorm** | 1098682 | {14 (nefazodone hydrochloride 100 MG Oral Tablet [Serzone]) / 14 (nefazodone hydrochloride 200 MG Oral Tablet [Serzone]) / 14 (nefazodone hydrochloride 50 MG Oral Tablet [Serzone]) } Pack [Serzone Starter Pack] |
| **RxNorm** | 1098709 | nefazodone hydrochloride 300 MG |
| **RxNorm** | 1098710 | nefazodone hydrochloride 300 MG Oral Tablet |
| **RxNorm** | 1098711 | nefazodone hydrochloride 25 MG |
| **RxNorm** | 1098712 | nefazodone hydrochloride 25 MG Oral Tablet |
| **RxNorm** | 1098713 | nefazodone hydrochloride 75 MG |
| **RxNorm** | 1098714 | nefazodone hydrochloride 75 MG Oral Tablet |
| **RxNorm** | 104824 | trazodone hydrochloride 150 MG Oral Tablet [Molipaxin] |
| **RxNorm** | 104825 | trazodone hydrochloride 150 MG Extended Release Oral Tablet [Molipaxin CR] |
| **RxNorm** | 104826 | trazodone hydrochloride 100 MG Oral Capsule [Molipaxin] |
| **RxNorm** | 104827 | trazodone hydrochloride 50 MG Oral Capsule [Molipaxin] |
| **RxNorm** | 104847 | fluvoxamine 50 MG Delayed Release Oral Tablet |
| **RxNorm** | 104848 | fluvoxamine 100 MG Delayed Release Oral Tablet |
| **RxNorm** | 104849 | fluoxetine 20 MG Oral Capsule [Prozac] |
| **RxNorm** | 104850 | fluoxetine 4 MG/ML Oral Solution [Prozac] |
| **RxNorm** | 114228 | Paxil |
| **RxNorm** | 1152130 | citalopram Oral Liquid Product |
| **RxNorm** | 1152131 | citalopram Oral Product |
| **RxNorm** | 1152132 | citalopram Pill |
| **RxNorm** | 1158193 | sertraline Oral Liquid Product |
| **RxNorm** | 1158194 | sertraline Oral Product |
| **RxNorm** | 1158195 | sertraline Pill |
| **RxNorm** | 1158404 | nefazodone Oral Product |
| **RxNorm** | 1158405 | nefazodone Pill |
| **RxNorm** | 1157677 | trazodone Injectable Product |
| **RxNorm** | 1157678 | trazodone Oral Liquid Product |
| **RxNorm** | 1157679 | trazodone Oral Product |
| **RxNorm** | 1157680 | trazodone Pill |
| **RxNorm** | 1166209 | Celexa Oral Liquid Product |
| **RxNorm** | 1166210 | Celexa Oral Product |
| **RxNorm** | 1166211 | Celexa Pill |
| **RxNorm** | 1164520 | escitalopram Oral Liquid Product |
| **RxNorm** | 1164521 | escitalopram Oral Product |
| **RxNorm** | 1164522 | escitalopram Pill |
| **RxNorm** | 1164136 | paroxetine Oral Liquid Product |
| **RxNorm** | 1164137 | paroxetine Oral Product |
| **RxNorm** | 1164138 | paroxetine Pill |

| | | |
|---|---|---|
| **RxNorm** | 1170894 | Cipramil Oral Product |
| **RxNorm** | 1170895 | Cipramil Pill |
| **RxNorm** | 1170912 | Citalopraam Oral Product |
| **RxNorm** | 1170913 | Citalopraam Pill |
| **RxNorm** | 1181637 | Molipaxin CR Oral Product |
| **RxNorm** | 1181638 | Molipaxin CR Pill |
| **RxNorm** | 1181639 | Molipaxin Oral Liquid Product |
| **RxNorm** | 1181640 | Molipaxin Oral Product |
| **RxNorm** | 1181641 | Molipaxin Pill |
| **RxNorm** | 1180777 | Serzone Oral Product |
| **RxNorm** | 1180778 | Serzone Pill |
| **RxNorm** | 1160833 | fluoxetine / olanzapine Oral Product |
| **RxNorm** | 1182066 | Luvox Oral Product |
| **RxNorm** | 1182067 | Luvox Pill |
| **RxNorm** | 1160834 | fluoxetine / olanzapine Pill |
| **RxNorm** | 1160835 | fluoxetine Oral Liquid Product |
| **RxNorm** | 1160836 | fluoxetine Oral Product |
| **RxNorm** | 1160837 | fluoxetine Pill |
| **RxNorm** | 1175211 | Desyrel Oral Product |
| **RxNorm** | 1175212 | Desyrel Pill |
| **RxNorm** | 1181164 | Oleptro Oral Product |
| **RxNorm** | 1181165 | Oleptro Pill |
| **RxNorm** | 1161460 | fluvoxamine Oral Product |
| **RxNorm** | 1161461 | fluvoxamine Pill |
| **RxNorm** | 1182485 | Prozac Oral Liquid Product |
| **RxNorm** | 1182486 | Prozac Oral Product |
| **RxNorm** | 1182487 | Prozac Pill |
| **RxNorm** | 1182920 | Paxil Oral Liquid Product |
| **RxNorm** | 1182921 | Paxil Oral Product |
| **RxNorm** | 1182922 | Paxil Pill |
| **RxNorm** | 1231897 | trazodone hydrochloride 150 MG [Molipaxin CR] |
| **RxNorm** | 1190110 | fluoxetine 60 MG Oral Tablet |
| **RxNorm** | 1185993 | Sarafem Oral Product |
| **RxNorm** | 1185994 | Sarafem Pill |
| **RxNorm** | 1186152 | Lexapro Oral Liquid Product |
| **RxNorm** | 1186153 | Lexapro Oral Product |
| **RxNorm** | 1186154 | Lexapro Pill |
| **RxNorm** | 1178223 | Pexeva Oral Product |
| **RxNorm** | 1178224 | Pexeva Pill |
| **RxNorm** | 1178280 | Rapiflux Oral Product |
| **RxNorm** | 1178281 | Rapiflux Pill |
| **RxNorm** | 1178362 | Selfemra Oral Product |
| **RxNorm** | 1178363 | Selfemra Pill |

| | | |
|---|---|---|
| **RxNorm** | 1185498 | Symbyax Oral Product |
| **RxNorm** | 1185499 | Symbyax Pill |
| **RxNorm** | 1188167 | Zoloft Oral Liquid Product |
| **RxNorm** | 1188168 | Zoloft Oral Product |
| **RxNorm** | 1188169 | Zoloft Pill |
| **RxNorm** | 1297094 | RECONCILE Chewable Product |
| **RxNorm** | 1177143 | RECONCILE Oral Product |
| **RxNorm** | 1177144 | RECONCILE Pill |
| **RxNorm** | 1295194 | fluoxetine Chewable Product |
| **RxNorm** | 1295242 | citalopram Disintegrating Oral Product |
| **RxNorm** | 141947 | trazodone hydrochloride 10 MG/ML Oral Solution [Molipaxin] |
| **RxNorm** | 1430121 | paroxetine Oral Capsule |
| **RxNorm** | 1430122 | paroxetine mesylate 7.5 MG Oral Capsule |
| **RxNorm** | 1430123 | Brisdelle |
| **RxNorm** | 1430125 | paroxetine Oral Capsule [Brisdelle] |
| **RxNorm** | 1430126 | Brisdelle Oral Product |
| **RxNorm** | 1430127 | Brisdelle Pill |
| **RxNorm** | 1430128 | paroxetine mesylate 7.5 MG Oral Capsule [Brisdelle] |
| **RxNorm** | 151260 | fluoxetine 60 MG Oral Capsule |
| **RxNorm** | 151261 | fluoxetine 60 MG Oral Capsule [Prozac] |
| **RxNorm** | 151516 | Cipramil |
| **RxNorm** | 151679 | Serzone |
| **RxNorm** | 153503 | citalopram 10 MG Oral Tablet [Cipramil] |
| **RxNorm** | 1738482 | paroxetine hydrochloride 10 MG |
| **RxNorm** | 1738483 | paroxetine hydrochloride 10 MG Oral Tablet |
| **RxNorm** | 1738484 | paroxetine hydrochloride 10 MG [Paxil] |
| **RxNorm** | 1738486 | paroxetine hydrochloride 12.5 MG |
| **RxNorm** | 1738488 | paroxetine hydrochloride 12.5 MG [Paxil] |
| **RxNorm** | 1738490 | paroxetine hydrochloride 2 MG/ML |
| **RxNorm** | 1738492 | paroxetine hydrochloride 2 MG/ML [Paxil] |
| **RxNorm** | 1738494 | paroxetine hydrochloride 20 MG |
| **RxNorm** | 1738495 | paroxetine hydrochloride 20 MG Oral Tablet |
| **RxNorm** | 1738496 | paroxetine hydrochloride 20 MG [Paxil] |
| **RxNorm** | 1738498 | paroxetine hydrochloride 25 MG |
| **RxNorm** | 1738500 | paroxetine hydrochloride 25 MG [Paxil] |
| **RxNorm** | 1738502 | paroxetine hydrochloride 30 MG |
| **RxNorm** | 1738503 | paroxetine hydrochloride 30 MG Oral Tablet |
| **RxNorm** | 1738504 | paroxetine hydrochloride 30 MG [Paxil] |
| **RxNorm** | 1738506 | paroxetine hydrochloride 37.5 MG |
| **RxNorm** | 1738508 | paroxetine hydrochloride 37.5 MG [Paxil] |
| **RxNorm** | 1738510 | paroxetine hydrochloride 40 MG |
| **RxNorm** | 1738511 | paroxetine hydrochloride 40 MG Oral Tablet |

| | | |
|---|---|---|
| **RxNorm** | 1738512 | paroxetine hydrochloride 40 MG [Paxil] |
| **RxNorm** | 1738514 | paroxetine mesylate 10 MG |
| **RxNorm** | 1738515 | paroxetine mesylate 10 MG Oral Tablet |
| **RxNorm** | 1738516 | paroxetine mesylate 10 MG [Pexeva] |
| **RxNorm** | 1738518 | paroxetine mesylate 20 MG |
| **RxNorm** | 1738519 | paroxetine mesylate 20 MG Oral Tablet |
| **RxNorm** | 1738520 | paroxetine mesylate 20 MG [Pexeva] |
| **RxNorm** | 1738522 | paroxetine mesylate 30 MG |
| **RxNorm** | 1738523 | paroxetine mesylate 30 MG Oral Tablet |
| **RxNorm** | 1738524 | paroxetine mesylate 30 MG [Pexeva] |
| **RxNorm** | 1738526 | paroxetine mesylate 40 MG |
| **RxNorm** | 1738527 | paroxetine mesylate 40 MG Oral Tablet |
| **RxNorm** | 1738528 | paroxetine mesylate 40 MG [Pexeva] |
| **RxNorm** | 1738562 | paroxetine mesylate 7.5 MG |
| **RxNorm** | 1738564 | paroxetine mesylate 7.5 MG [Brisdelle] |
| **RxNorm** | 1738803 | 24 HR paroxetine hydrochloride 12.5 MG Extended Release Oral Tablet |
| **RxNorm** | 1738804 | 24 HR paroxetine hydrochloride 12.5 MG Extended Release Oral Tablet [Paxil] |
| **RxNorm** | 1738805 | 24 HR paroxetine hydrochloride 25 MG Extended Release Oral Tablet |
| **RxNorm** | 1738806 | 24 HR paroxetine hydrochloride 25 MG Extended Release Oral Tablet [Paxil] |
| **RxNorm** | 1738807 | 24 HR paroxetine hydrochloride 37.5 MG Extended Release Oral Tablet |
| **RxNorm** | 1738808 | 24 HR paroxetine hydrochloride 37.5 MG Extended Release Oral Tablet [Paxil] |
| **RxNorm** | 200371 | citalopram 20 MG Oral Tablet |
| **RxNorm** | 199990 | sertraline 25 MG Oral Capsule |
| **RxNorm** | 204192 | Desyrel |
| **RxNorm** | 205535 | fluoxetine 10 MG Oral Capsule [Prozac] |
| **RxNorm** | 207350 | paroxetine hydrochloride 30 MG Oral Tablet [Paxil] |
| **RxNorm** | 207349 | paroxetine hydrochloride 20 MG Oral Tablet [Paxil] |
| **RxNorm** | 208149 | sertraline 100 MG Oral Tablet [Zoloft] |
| **RxNorm** | 208161 | sertraline 50 MG Oral Tablet [Zoloft] |
| **RxNorm** | 211699 | paroxetine hydrochloride 10 MG Oral Tablet [Paxil] |
| **RxNorm** | 211700 | paroxetine hydrochloride 40 MG Oral Tablet [Paxil] |
| **RxNorm** | 212233 | sertraline 25 MG Oral Tablet [Zoloft] |
| **RxNorm** | 213291 | paroxetine hydrochloride 2 MG/ML Oral Suspension [Paxil] |
| **RxNorm** | 213344 | citalopram 20 MG Oral Tablet [Celexa] |
| **RxNorm** | 213345 | citalopram 40 MG Oral Tablet [Celexa] |
| **RxNorm** | 215928 | Celexa |
| **RxNorm** | 226376 | citalopram 40 MG Oral Tablet [Cipramil] |
| **RxNorm** | 225514 | Molipaxin CR |

| | | |
|---|---|---|
| **RxNorm** | 248642 | fluoxetine 20 MG Oral Tablet |
| **RxNorm** | 248097 | sertraline 50 MG Oral Capsule |
| **RxNorm** | 248098 | sertraline 100 MG Oral Capsule |
| **RxNorm** | 252271 | fluoxetine 3 MG Oral Tablet |
| **RxNorm** | 252272 | fluoxetine 6 MG Oral Tablet |
| **RxNorm** | 252440 | fluoxetine 12 MG Oral Tablet |
| **RxNorm** | 284591 | citalopram 10 MG Oral Tablet [Celexa] |
| **RxNorm** | 261282 | fluoxetine 10 MG Oral Tablet [Prozac] |
| **RxNorm** | 261287 | fluoxetine 40 MG Oral Capsule [Prozac] |
| **RxNorm** | 251200 | sertraline 200 MG Oral Tablet |
| **RxNorm** | 251201 | sertraline 200 MG Oral Capsule |
| **RxNorm** | 261342 | citalopram 2 MG/ML Oral Solution [Celexa] |
| **RxNorm** | 251494 | trazodone 75 MG Oral Tablet |
| **RxNorm** | 310384 | fluoxetine 10 MG Oral Capsule |
| **RxNorm** | 310385 | fluoxetine 20 MG Oral Capsule |
| **RxNorm** | 310386 | fluoxetine 4 MG/ML Oral Solution |
| **RxNorm** | 309313 | citalopram 2 MG/ML Oral Solution |
| **RxNorm** | 309314 | citalopram 40 MG Oral Tablet |
| **RxNorm** | 281674 | Molipaxin |
| **RxNorm** | 283672 | citalopram 10 MG Oral Tablet |
| **RxNorm** | 312938 | sertraline 100 MG Oral Tablet |
| **RxNorm** | 312940 | sertraline 25 MG Oral Tablet |
| **RxNorm** | 312941 | sertraline 50 MG Oral Tablet |
| **RxNorm** | 284196 | fluoxetine 90 MG Extended Release Oral Tablet |
| **RxNorm** | 312242 | paroxetine hydrochloride 2 MG/ML Oral Suspension |
| **RxNorm** | 313989 | fluoxetine 40 MG Oral Capsule |
| **RxNorm** | 313990 | fluoxetine 10 MG Oral Tablet |
| **RxNorm** | 313995 | fluoxetine 90 MG Delayed Release Oral Capsule |
| **RxNorm** | 315951 | fluoxetine 10 MG |
| **RxNorm** | 315952 | fluoxetine 20 MG |
| **RxNorm** | 315953 | fluoxetine 4 MG/ML |
| **RxNorm** | 329212 | sertraline 25 MG |
| **RxNorm** | 329444 | citalopram 20 MG |
| **RxNorm** | 329445 | citalopram 40 MG |
| **RxNorm** | 331559 | fluoxetine 90 MG |
| **RxNorm** | 331567 | citalopram 10 MG |
| **RxNorm** | 336689 | fluoxetine 6 MG |
| **RxNorm** | 330309 | citalopram 2 MG/ML |
| **RxNorm** | 330341 | fluoxetine 40 MG |
| **RxNorm** | 336997 | trazodone 75 MG |
| **RxNorm** | 349332 | escitalopram 10 MG Oral Tablet |
| **RxNorm** | 328268 | fluvoxamine 50 MG |
| **RxNorm** | 328269 | fluvoxamine 100 MG |

| | | |
|---|---|---|
| **RxNorm** | 352004 | fluoxetine 20 MG Oral Tablet [Rapiflux] |
| **RxNorm** | 352272 | escitalopram 10 MG Oral Tablet [Lexapro] |
| **RxNorm** | 352273 | escitalopram 20 MG Oral Tablet [Lexapro] |
| **RxNorm** | 334494 | sertraline 200 MG |
| **RxNorm** | 351044 | escitalopram 10 MG |
| **RxNorm** | 335767 | trazodone 10 MG/ML |
| **RxNorm** | 351249 | escitalopram 5 MG Oral Tablet |
| **RxNorm** | 351250 | escitalopram 20 MG Oral Tablet |
| **RxNorm** | 328670 | sertraline 100 MG |
| **RxNorm** | 328671 | sertraline 50 MG |
| **RxNorm** | 352741 | Lexapro |
| **RxNorm** | 351285 | escitalopram 1 MG/ML Oral Solution |
| **RxNorm** | 334800 | fluoxetine 60 MG |
| **RxNorm** | 334801 | fluoxetine 12 MG |
| **RxNorm** | 334802 | fluoxetine 3 MG |
| **RxNorm** | 366082 | paroxetine Oral Suspension [Paxil] |
| **RxNorm** | 352940 | Rapiflux |
| **RxNorm** | 367670 | paroxetine Oral Tablet [Paxil] |
| **RxNorm** | 353382 | escitalopram 5 MG |
| **RxNorm** | 353383 | escitalopram 20 MG |
| **RxNorm** | 353398 | escitalopram 1 MG/ML |
| **RxNorm** | 366526 | trazodone Oral Capsule [Molipaxin] |
| **RxNorm** | 363958 | fluoxetine Oral Solution [Prozac] |
| **RxNorm** | 368139 | citalopram Oral Tablet [Celexa] |
| **RxNorm** | 364573 | sertraline Oral Solution [Zoloft] |
| **RxNorm** | 364574 | citalopram Oral Solution [Celexa] |
| **RxNorm** | 368226 | fluoxetine Oral Tablet [Prozac] |
| **RxNorm** | 368306 | nefazodone Oral Tablet [Serzone] |
| **RxNorm** | 368413 | sertraline Oral Tablet [Zoloft] |
| **RxNorm** | 368436 | fluvoxamine Oral Tablet [Luvox] |
| **RxNorm** | 368503 | citalopram Oral Tablet [Cipramil] |
| **RxNorm** | 368571 | escitalopram Oral Tablet [Lexapro] |
| **RxNorm** | 369258 | trazodone Oral Tablet [Desyrel] |
| **RxNorm** | 369442 | trazodone Oral Tablet [Molipaxin] |
| **RxNorm** | 369487 | fluoxetine Oral Tablet [Rapiflux] |
| **RxNorm** | 370119 | trazodone Extended Release Oral Tablet [Molipaxin CR] |
| **RxNorm** | 371532 | citalopram Oral Tablet |
| **RxNorm** | 374620 | citalopram Oral Solution |
| **RxNorm** | 374622 | sertraline Oral Solution |
| **RxNorm** | 374650 | fluoxetine Extended Release Oral Tablet |
| **RxNorm** | 372231 | fluoxetine Oral Capsule |
| **RxNorm** | 372232 | fluoxetine Oral Solution |
| **RxNorm** | 372233 | fluoxetine Oral Tablet |

| | | |
|---|---|---|
| **RxNorm** | 372256 | fluvoxamine Oral Tablet |
| **RxNorm** | 380863 | trazodone Oral Solution [Molipaxin] |
| **RxNorm** | 389202 | citalopram 40 MG/ML Oral Solution |
| **RxNorm** | 373867 | sertraline Oral Capsule |
| **RxNorm** | 373868 | sertraline Oral Tablet |
| **RxNorm** | 376314 | trazodone Extended Release Oral Tablet |
| **RxNorm** | 378618 | fluoxetine Delayed Release Oral Capsule |
| **RxNorm** | 378695 | escitalopram Oral Tablet |
| **RxNorm** | 378734 | escitalopram Oral Solution |
| **RxNorm** | 373012 | nefazodone Oral Tablet |
| **RxNorm** | 374183 | trazodone Oral Capsule |
| **RxNorm** | 374184 | trazodone Oral Solution |
| **RxNorm** | 374185 | trazodone Oral Tablet |
| **RxNorm** | 378925 | paroxetine Oral Tablet |
| **RxNorm** | 393457 | citalopram 40 MG/ML |
| **RxNorm** | 410584 | sertraline 150 MG Oral Capsule |
| **RxNorm** | 373245 | paroxetine Oral Suspension |
| **RxNorm** | 405343 | Symbyax |
| **RxNorm** | 411880 | trazodone 25 MG Oral Capsule |
| **RxNorm** | 413145 | trazodone 10 MG/ML Injectable Solution |
| **RxNorm** | 385706 | fluvoxamine Delayed Release Oral Tablet |
| **RxNorm** | 406022 | fluoxetine 25 MG |
| **RxNorm** | 406024 | fluoxetine / olanzapine Oral Capsule |
| **RxNorm** | 406026 | fluoxetine 50 MG |
| **RxNorm** | 406125 | escitalopram Oral Solution [Lexapro] |
| **RxNorm** | 406413 | fluoxetine 40 MG/ML Oral Solution |
| **RxNorm** | 42687 | Luvox |
| **RxNorm** | 433731 | fluoxetine 20 MG Delayed Release Oral Tablet |
| **RxNorm** | 403969 | fluoxetine 25 MG / olanzapine 6 MG Oral Capsule |
| **RxNorm** | 403970 | fluoxetine 25 MG / olanzapine 12 MG Oral Capsule |
| **RxNorm** | 403971 | fluoxetine 50 MG / olanzapine 12 MG Oral Capsule |
| **RxNorm** | 403972 | fluoxetine 50 MG / olanzapine 6 MG Oral Capsule |
| **RxNorm** | 404408 | escitalopram 5 MG Oral Tablet [Lexapro] |
| **RxNorm** | 404420 | escitalopram 1 MG/ML Oral Solution [Lexapro] |
| **RxNorm** | 439360 | trazodone 25 MG |
| **RxNorm** | 439755 | fluoxetine Delayed Release Oral Tablet |
| **RxNorm** | 540845 | Citalopraam |
| **RxNorm** | 540846 | citalopram 20 MG [Citalopraam] |
| **RxNorm** | 540847 | citalopram Oral Tablet [Citalopraam] |
| **RxNorm** | 540848 | citalopram 20 MG Oral Tablet [Citalopraam] |
| **RxNorm** | 541357 | fluoxetine 40 MG Oral Tablet |
| **RxNorm** | 541517 | fluoxetine 20 MG/ML |
| **RxNorm** | 541518 | fluoxetine 20 MG/ML Oral Solution |

| | | |
|---|---|---|
| **RxNorm** | 541519 | fluoxetine 20 MG/ML [Prozac] |
| **RxNorm** | 541520 | fluoxetine 20 MG/ML Oral Solution [Prozac] |
| **RxNorm** | 541657 | Pexeva |
| **RxNorm** | 541659 | paroxetine Oral Tablet [Pexeva] |
| **RxNorm** | 541660 | paroxetine mesylate 10 MG Oral Tablet [Pexeva] |
| **RxNorm** | 541662 | paroxetine mesylate 20 MG Oral Tablet [Pexeva] |
| **RxNorm** | 541664 | paroxetine mesylate 30 MG Oral Tablet [Pexeva] |
| **RxNorm** | 541666 | paroxetine mesylate 40 MG Oral Tablet [Pexeva] |
| **RxNorm** | 451487 | sertraline 150 MG |
| **RxNorm** | 452327 | trazodone Injectable Solution |
| **RxNorm** | 484801 | fluoxetine 50 MG / olanzapine 60 MG Oral Capsule |
| **RxNorm** | 539701 | fluoxetine 90 MG [Prozac] |
| **RxNorm** | 564629 | citalopram 20 MG [Cipramil] |
| **RxNorm** | 452715 | fluoxetine 40 MG/ML |
| **RxNorm** | 564960 | fluoxetine 60 MG [Prozac] |
| **RxNorm** | 565289 | citalopram 10 MG [Cipramil] |
| **RxNorm** | 563783 | fluoxetine 20 MG [Prozac] |
| **RxNorm** | 563784 | fluoxetine 4 MG/ML [Prozac] |
| **RxNorm** | 566436 | fluoxetine 10 MG [Prozac] |
| **RxNorm** | 568875 | sertraline 100 MG [Zoloft] |
| **RxNorm** | 568885 | sertraline 50 MG [Zoloft] |
| **RxNorm** | 562789 | paroxetine Extended Release Oral Tablet |
| **RxNorm** | 562790 | paroxetine hydrochloride 12.5 MG Extended Release Oral Tablet |
| **RxNorm** | 562791 | paroxetine hydrochloride 25 MG Extended Release Oral Tablet |
| **RxNorm** | 562815 | paroxetine hydrochloride 37.5 MG Extended Release Oral Tablet |
| **RxNorm** | 572455 | sertraline 25 MG [Zoloft] |
| **RxNorm** | 575018 | citalopram 10 MG [Celexa] |
| **RxNorm** | 575748 | fluoxetine 20 MG [Rapiflux] |
| **RxNorm** | 575972 | escitalopram 10 MG [Lexapro] |
| **RxNorm** | 575973 | escitalopram 20 MG [Lexapro] |
| **RxNorm** | 598031 | fluoxetine Delayed Release Oral Capsule [Prozac] |
| **RxNorm** | 598032 | fluoxetine 90 MG Delayed Release Oral Capsule [Prozac] |
| **RxNorm** | 573240 | citalopram 20 MG [Celexa] |
| **RxNorm** | 573241 | citalopram 40 MG [Celexa] |
| **RxNorm** | 576370 | escitalopram 5 MG [Lexapro] |
| **RxNorm** | 576381 | escitalopram 1 MG/ML [Lexapro] |
| **RxNorm** | 58827 | Prozac |
| **RxNorm** | 611247 | fluoxetine / olanzapine |
| **RxNorm** | 573976 | citalopram 40 MG [Cipramil] |
| **RxNorm** | 574512 | fluoxetine 40 MG [Prozac] |

| | | |
|---|---|---|
| **RxNorm** | 574562 | citalopram 2 MG/ML [Celexa] |
| **RxNorm** | 645288 | citalopram Disintegrating Oral Tablet |
| **RxNorm** | 645289 | citalopram 10 MG Disintegrating Oral Tablet |
| **RxNorm** | 645290 | citalopram 20 MG Disintegrating Oral Tablet |
| **RxNorm** | 645291 | citalopram 40 MG Disintegrating Oral Tablet |
| **RxNorm** | 647323 | fluoxetine 10 MG [Sarafem] |
| **RxNorm** | 647324 | fluoxetine Oral Capsule [Sarafem] |
| **RxNorm** | 647325 | fluoxetine 10 MG Oral Capsule [Sarafem] |
| **RxNorm** | 639464 | Sarafem |
| **RxNorm** | 647441 | fluoxetine 20 MG [Sarafem] |
| **RxNorm** | 647442 | fluoxetine 20 MG Oral Capsule [Sarafem] |
| **RxNorm** | 647554 | fluoxetine Oral Tablet [Sarafem] |
| **RxNorm** | 647555 | fluoxetine 10 MG Oral Tablet [Sarafem] |
| **RxNorm** | 647556 | fluoxetine 20 MG Oral Tablet [Sarafem] |
| **RxNorm** | 631739 | citalopram 3 MG/ML |
| **RxNorm** | 631740 | citalopram 3 MG/ML Oral Solution |
| **RxNorm** | 729704 | RECONCILE |
| **RxNorm** | 730439 | citalopram Oral Capsule |
| **RxNorm** | 730440 | citalopram 10 MG Oral Capsule |
| **RxNorm** | 730441 | citalopram 20 MG Oral Capsule |
| **RxNorm** | 730442 | citalopram 40 MG Oral Capsule |
| **RxNorm** | 721787 | fluoxetine 25 MG / olanzapine 3 MG Oral Capsule |
| **RxNorm** | 725062 | fluoxetine 25 MG / olanzapine 6 MG [Symbyax] |
| **RxNorm** | 725063 | fluoxetine / olanzapine Oral Capsule [Symbyax] |
| **RxNorm** | 790408 | fluvoxamine Extended Release Oral Capsule |
| **RxNorm** | 790880 | fluvoxamine Extended Release Oral Capsule [Luvox] |
| **RxNorm** | 725064 | fluoxetine 25 MG / olanzapine 6 MG Oral Capsule [Symbyax] |
| **RxNorm** | 725066 | fluoxetine 50 MG / olanzapine 12 MG [Symbyax] |
| **RxNorm** | 725068 | fluoxetine 50 MG / olanzapine 12 MG Oral Capsule [Symbyax] |
| **RxNorm** | 725070 | fluoxetine 50 MG / olanzapine 6 MG [Symbyax] |
| **RxNorm** | 725072 | fluoxetine 50 MG / olanzapine 6 MG Oral Capsule [Symbyax] |
| **RxNorm** | 725074 | fluoxetine 25 MG / olanzapine 12 MG [Symbyax] |
| **RxNorm** | 725076 | fluoxetine 25 MG / olanzapine 12 MG Oral Capsule [Symbyax] |
| **RxNorm** | 725078 | fluoxetine 25 MG / olanzapine 3 MG [Symbyax] |
| **RxNorm** | 725080 | fluoxetine 25 MG / olanzapine 3 MG Oral Capsule [Symbyax] |
| **RxNorm** | 799023 | Selfemra |
| **RxNorm** | 799024 | fluoxetine 10 MG [Selfemra] |
| **RxNorm** | 799025 | fluoxetine Oral Capsule [Selfemra] |
| **RxNorm** | 799026 | fluoxetine 10 MG Oral Capsule [Selfemra] |

| | | |
|---|---|---|
| **RxNorm** | 799058 | fluoxetine 20 MG [Selfemra] |
| **RxNorm** | 799059 | fluoxetine 20 MG Oral Capsule [Selfemra] |
| **RxNorm** | 794947 | sertraline 150 MG Oral Tablet |
| **RxNorm** | 826096 | fluoxetine hydrochloride 8 MG |
| **RxNorm** | 826097 | fluoxetine Chewable Tablet |
| **RxNorm** | 826098 | fluoxetine hydrochloride 8 MG Chewable Tablet |
| **RxNorm** | 826099 | fluoxetine hydrochloride 8 MG [RECONCILE] |
| **RxNorm** | 826100 | fluoxetine Chewable Tablet [RECONCILE] |
| **RxNorm** | 826101 | fluoxetine hydrochloride 8 MG Chewable Tablet [RECONCILE] |
| **RxNorm** | 826102 | fluoxetine hydrochloride 64 MG |
| **RxNorm** | 826103 | fluoxetine hydrochloride 64 MG Chewable Tablet |
| **RxNorm** | 826104 | fluoxetine hydrochloride 64 MG [RECONCILE] |
| **RxNorm** | 826105 | fluoxetine hydrochloride 64 MG Chewable Tablet [RECONCILE] |
| **RxNorm** | 82728 | Zoloft |
| **RxNorm** | 828445 | paroxetine Extended Release Oral Tablet [Paxil] |
| **RxNorm** | 828446 | paroxetine hydrochloride 12.5 MG Extended Release Oral Tablet [Paxil] |
| **RxNorm** | 828448 | paroxetine hydrochloride 25 MG Extended Release Oral Tablet [Paxil] |
| **RxNorm** | 828451 | paroxetine hydrochloride 37.5 MG Extended Release Oral Tablet [Paxil] |
| **RxNorm** | 803292 | fluoxetine 15 MG |
| **RxNorm** | 803293 | fluoxetine 15 MG Oral Tablet |
| **RxNorm** | 803294 | fluoxetine 15 MG [Sarafem] |
| **RxNorm** | 803295 | fluoxetine 15 MG Oral Tablet [Sarafem] |
| **RxNorm** | 856363 | trazodone hydrochloride 150 MG |
| **RxNorm** | 856364 | trazodone hydrochloride 150 MG Oral Tablet |
| **RxNorm** | 856365 | trazodone hydrochloride 150 MG [Desyrel] |
| **RxNorm** | 856366 | trazodone hydrochloride 150 MG Oral Tablet [Desyrel] |
| **RxNorm** | 856368 | trazodone hydrochloride 300 MG |
| **RxNorm** | 856369 | trazodone hydrochloride 300 MG Oral Tablet |
| **RxNorm** | 856370 | trazodone hydrochloride 300 MG [Desyrel] |
| **RxNorm** | 856371 | trazodone hydrochloride 300 MG Oral Tablet [Desyrel] |
| **RxNorm** | 856372 | trazodone hydrochloride 100 MG |
| **RxNorm** | 856373 | trazodone hydrochloride 100 MG Oral Tablet |
| **RxNorm** | 856374 | trazodone hydrochloride 100 MG [Desyrel] |
| **RxNorm** | 856375 | trazodone hydrochloride 100 MG Oral Tablet [Desyrel] |
| **RxNorm** | 856376 | trazodone hydrochloride 50 MG |
| **RxNorm** | 856377 | trazodone hydrochloride 50 MG Oral Tablet |
| **RxNorm** | 856378 | trazodone hydrochloride 50 MG [Desyrel] |
| **RxNorm** | 856379 | trazodone hydrochloride 50 MG Oral Tablet [Desyrel] |
| **RxNorm** | 856380 | trazodone hydrochloride 10 MG/ML |

| | | |
|---|---|---|
| **RxNorm** | 856381 | trazodone hydrochloride 10 MG/ML Oral Solution |
| **RxNorm** | 856382 | trazodone hydrochloride 100 MG Oral Capsule |
| **RxNorm** | 856383 | trazodone hydrochloride 50 MG Oral Capsule |
| **RxNorm** | 875910 | trazodone hydrochloride 100 MG [Molipaxin] |
| **RxNorm** | 875911 | trazodone hydrochloride 50 MG [Molipaxin] |
| **RxNorm** | 875938 | trazodone hydrochloride 10 MG/ML [Molipaxin] |
| **RxNorm** | 876027 | trazodone hydrochloride 150 MG [Molipaxin] |
| **RxNorm** | 861063 | sertraline 20 MG/ML |
| **RxNorm** | 861064 | sertraline 20 MG/ML Oral Solution |
| **RxNorm** | 861065 | sertraline 20 MG/ML [Zoloft] |
| **RxNorm** | 861066 | sertraline 20 MG/ML Oral Solution [Zoloft] |
| **RxNorm** | 898705 | trazodone hydrochloride 300 MG [Oleptro] |
| **RxNorm** | 898706 | 24 HR trazodone hydrochloride 300 MG Extended Release Oral Tablet [Oleptro] |
| **RxNorm** | 898707 | trazodone hydrochloride 300 MG Extended Release Oral Tablet |
| **RxNorm** | 898708 | trazodone hydrochloride 300 MG Extended Release Oral Tablet [Oleptro] |
| **RxNorm** | 902713 | fluoxetine hydrochloride 16 MG |
| **RxNorm** | 902714 | fluoxetine hydrochloride 16 MG Chewable Tablet |
| **RxNorm** | 902715 | fluoxetine hydrochloride 16 MG [RECONCILE] |
| **RxNorm** | 902716 | fluoxetine hydrochloride 16 MG Chewable Tablet [RECONCILE] |
| **RxNorm** | 902717 | fluoxetine hydrochloride 32 MG |
| **RxNorm** | 902718 | fluoxetine hydrochloride 32 MG Chewable Tablet |
| **RxNorm** | 902719 | fluoxetine hydrochloride 32 MG [RECONCILE] |
| **RxNorm** | 902720 | fluoxetine hydrochloride 32 MG Chewable Tablet [RECONCILE] |
| **RxNorm** | 903872 | fluvoxamine maleate 100 MG |
| **RxNorm** | 903873 | 24 HR fluvoxamine maleate 100 MG Extended Release Oral Capsule |
| **RxNorm** | 903874 | fluvoxamine maleate 100 MG [Luvox] |
| **RxNorm** | 903875 | 24 HR fluvoxamine maleate 100 MG Extended Release Oral Capsule [Luvox] |
| **RxNorm** | 903876 | fluvoxamine maleate 100 MG Extended Release Oral Capsule |
| **RxNorm** | 903877 | fluvoxamine maleate 100 MG Extended Release Oral Capsule [Luvox] |
| **RxNorm** | 903878 | fluvoxamine maleate 150 MG |
| **RxNorm** | 903879 | 24 HR fluvoxamine maleate 150 MG Extended Release Oral Capsule |
| **RxNorm** | 903880 | fluvoxamine maleate 150 MG [Luvox] |
| **RxNorm** | 903881 | 24 HR fluvoxamine maleate 150 MG Extended Release Oral Capsule [Luvox] |

| | | |
|---|---|---|
| **RxNorm** | 903882 | fluvoxamine maleate 150 MG Extended Release Oral Capsule |
| **RxNorm** | 903883 | fluvoxamine maleate 150 MG Extended Release Oral Capsule [Luvox] |
| **RxNorm** | 903884 | fluvoxamine maleate 100 MG Oral Tablet |
| **RxNorm** | 903885 | fluvoxamine maleate 100 MG Oral Tablet [Luvox] |
| **RxNorm** | 903886 | fluvoxamine maleate 25 MG |
| **RxNorm** | 903887 | fluvoxamine maleate 25 MG Oral Tablet |
| **RxNorm** | 903888 | fluvoxamine maleate 25 MG [Luvox] |
| **RxNorm** | 903889 | fluvoxamine maleate 25 MG Oral Tablet [Luvox] |
| **RxNorm** | 903890 | fluvoxamine maleate 50 MG |
| **RxNorm** | 903891 | fluvoxamine maleate 50 MG Oral Tablet |
| **RxNorm** | 903892 | fluvoxamine maleate 50 MG [Luvox] |
| **RxNorm** | 903893 | fluvoxamine maleate 50 MG Oral Tablet [Luvox] |
| **RxNorm** | 898697 | 24 HR trazodone hydrochloride 150 MG Extended Release Oral Tablet |
| **RxNorm** | 898698 | Oleptro |
| **RxNorm** | 898699 | trazodone hydrochloride 150 MG [Oleptro] |
| **RxNorm** | 898700 | trazodone Extended Release Oral Tablet [Oleptro] |
| **RxNorm** | 898701 | 24 HR trazodone hydrochloride 150 MG Extended Release Oral Tablet [Oleptro] |
| **RxNorm** | 898702 | trazodone hydrochloride 150 MG Extended Release Oral Tablet |
| **RxNorm** | 898703 | trazodone hydrochloride 150 MG Extended Release Oral Tablet [Oleptro] |
| **RxNorm** | 898704 | 24 HR trazodone hydrochloride 300 MG Extended Release Oral Tablet |
| **RxNorm** | 93904 | fluoxetine Oral Capsule [Prozac] |
| **RxNorm** | 825431 | Sertraline 0.2 MG/ML Oral Solution [Zoloft] |
| **RxNorm** | 198299 | Trazodone 150 MG Oral Tablet |
| **RxNorm** | 313449 | Trazodone 50 MG Oral Tablet |
| **RxNorm** | 328688 | Trazodone 150 MG |
| **RxNorm** | 328854 | Trazodone 300 MG |
| **RxNorm** | 316329 | nefazodone 100 MG |
| **RxNorm** | 313994 | Fluvoxamine 50 MG Oral Tablet |
| **RxNorm** | 579898 | Fluoxetine 90 MG Extended Release Oral Capsule |
| **RxNorm** | 211995 | Fluvoxamine 25 MG Oral Tablet [Luvox] |
| **RxNorm** | 316331 | nefazodone 250 MG |
| **RxNorm** | 317435 | nefazodone 200 MG |
| **RxNorm** | 252439 | nefazodone 75 MG Oral Tablet |
| **RxNorm** | 311923 | nefazodone 50 MG Oral Tablet |
| **RxNorm** | 312241 | Paroxetine 10 MG Oral Tablet |
| **RxNorm** | 312244 | Paroxetine 40 MG Oral Tablet |
| **RxNorm** | 198298 | Trazodone 100 MG Oral Tablet |

| | | |
|---|---|---|
| **RxNorm** | 252438 | nefazodone 25 MG Oral Tablet |
| **RxNorm** | 562162 | Paroxetine 2 MG/ML Oral Solution |
| **RxNorm** | 312243 | Paroxetine 30 MG Oral Tablet |
| **RxNorm** | 314199 | Paroxetine 20 MG Oral Tablet |
| **RxNorm** | 316446 | Paroxetine 40 MG |
| **RxNorm** | 329177 | Fluvoxamine 25 MG |
| **RxNorm** | 336761 | nefazodone 75 MG |
| **RxNorm** | 316445 | Paroxetine 30 MG |
| **RxNorm** | 316444 | Paroxetine 2 MG/ML |
| **RxNorm** | 316443 | Paroxetine 10 MG |
| **RxNorm** | 317659 | Paroxetine 20 MG |
| **RxNorm** | 207114 | nefazodone 100 MG Oral Tablet [Serzone] |
| **RxNorm** | 207116 | nefazodone 200 MG Oral Tablet [Serzone] |
| **RxNorm** | 566570 | Fluvoxamine 100 MG [Luvox] |
| **RxNorm** | 350634 | Paroxetine 37.5 MG |
| **RxNorm** | 351962 | Fluoxetine 90 MG Enteric Coated Capsule [Prozac Weekly] |
| **RxNorm** | 207115 | nefazodone 150 MG Oral Tablet [Serzone] |
| **RxNorm** | 207117 | nefazodone 250 MG Oral Tablet [Serzone] |
| **RxNorm** | 260419 | Trazodone 300 MG Oral Tablet |
| **RxNorm** | 104828 | Trazodone 10 MG/ML Oral Solution |
| **RxNorm** | 351128 | nefazodone 300 MG Oral Tablet |
| **RxNorm** | 353351 | nefazodone 300 MG |
| **RxNorm** | 567906 | nefazodone 150 MG [Serzone] |
| **RxNorm** | 567905 | nefazodone 100 MG [Serzone] |
| **RxNorm** | 567907 | nefazodone 200 MG [Serzone] |
| **RxNorm** | 569029 | Trazodone 150 MG [Desyrel Dividose] |
| **RxNorm** | 208315 | Trazodone 100 MG Oral Tablet [Desyrel] |
| **RxNorm** | 569023 | Trazodone 100 MG [Desyrel] |
| **RxNorm** | 569021 | Trazodone 50 MG [Desyrel] |
| **RxNorm** | 208313 | Trazodone 50 MG Oral Tablet [Desyrel] |
| **RxNorm** | 208322 | Trazodone 150 MG Oral Tablet [Desyrel Dividose] |
| **RxNorm** | 567908 | nefazodone 250 MG [Serzone] |
| **RxNorm** | 572042 | Paroxetine 10 MG [Paxil] |
| **RxNorm** | 572043 | Paroxetine 40 MG [Paxil] |
| **RxNorm** | 573029 | nefazodone 50 MG [Serzone] |
| **RxNorm** | 573201 | Paroxetine 2 MG/ML [Paxil] |
| **RxNorm** | 541661 | Paroxetine 20 MG [Pexeva] |
| **RxNorm** | 541665 | Paroxetine 40 MG [Pexeva] |
| **RxNorm** | 541658 | Paroxetine 10 MG [Pexeva] |
| **RxNorm** | 541663 | Paroxetine 30 MG [Pexeva] |
| **RxNorm** | 209218 | Trazodone 300 MG Oral Tablet [Desyrel Dividose] |
| **RxNorm** | 393240 | Trazodone 50 MG Oral Capsule |
| **RxNorm** | 568123 | Paroxetine 20 MG [Paxil] |

| | | |
|---|---|---|
| **RxNorm** | 579900 | Fluoxetine 90 MG Extended Release Oral Capsule [Prozac Weekly] |
| **RxNorm** | 828450 | Paroxetine 37.5 MG [Paxil] |
| **RxNorm** | 828444 | Paroxetine 12.5 MG [Paxil] |
| **RxNorm** | 828447 | Paroxetine 25 MG [Paxil] |
| **RxNorm** | 568124 | Paroxetine 30 MG [Paxil] |
| **RxNorm** | 844753 | Paroxetine 25 MG Extended Release Oral Capsule |
| **RxNorm** | 583058 | Fluoxetine 90 MG Extended Release Oral Capsule [Prozac] |
| **RxNorm** | 150817 | Trazodone 100 MG Oral Capsule |
| **RxNorm** | 213100 | nefazodone 50 MG Oral Tablet [Serzone] |
| **RxNorm** | 153430 | Paroxetine 2 MG/ML Oral Solution [Seroxat] |
| **RxNorm** | 685570 | Fluoxetine 90 MG [Prozac Weekly] |
| **RxNorm** | 566569 | Fluvoxamine 50 MG [Luvox] |
| **RxNorm** | 310408 | Fluvoxamine 25 MG Oral Tablet |
| **RxNorm** | 310407 | Fluvoxamine 100 MG Oral Tablet |
| **RxNorm** | 572312 | Fluvoxamine 25 MG [Luvox] |
| **RxNorm** | 316330 | nefazodone 150 MG |
| **RxNorm** | 330509 | Trazodone 50 MG |
| **RxNorm** | 330510 | Trazodone 100 MG |
| **RxNorm** | 204454 | nefazodone 150 MG Oral Tablet |
| **RxNorm** | 204455 | nefazodone 200 MG Oral Tablet |
| **RxNorm** | 330794 | nefazodone 50 MG |
| **RxNorm** | 539702 | Fluoxetine 90 MG Oral Capsule [Prozac] |
| **RxNorm** | 539700 | Fluoxetine 90 MG Oral Capsule |
| **RxNorm** | 569845 | Trazodone 300 MG [Desyrel Dividose] |
| **RxNorm** | 790883 | 24 HR Fluvoxamine 150 MG Extended Release Capsule [Luvox] |
| **RxNorm** | 790410 | Fluvoxamine 150 MG |
| **RxNorm** | 250531 | Trazodone 150 MG Extended Release Tablet |
| **RxNorm** | 790409 | 24 HR Fluvoxamine 100 MG Extended Release Capsule |
| **RxNorm** | 790411 | 24 HR Fluvoxamine 150 MG Extended Release Capsule |
| **RxNorm** | 204453 | nefazodone 100 MG Oral Tablet |
| **RxNorm** | 790882 | Fluvoxamine 150 MG [Luvox] |
| **RxNorm** | 763194 | {14 (nefazodone 100 MG Oral Tablet) / 14 (nefazodone 200 MG Oral Tablet) / 14 (nefazodone 50 MG Oral Tablet) } Pack |
| **RxNorm** | 204456 | nefazodone 250 MG Oral Tablet |
| **RxNorm** | 790881 | 24 HR Fluvoxamine 100 MG Extended Release Capsule [Luvox] |
| **RxNorm** | 252528 | 24 HR Paroxetine 25 MG Extended Release Oral Capsule |
| **RxNorm** | 563774 | Trazodone 100 MG [Molipaxin] |
| **RxNorm** | 563775 | Trazodone 50 MG [Molipaxin] |
| **RxNorm** | 563773 | Trazodone 150 MG [Molipaxin CR] |
| **RxNorm** | 563772 | Trazodone 150 MG [Molipaxin] |

| | | |
|---|---|---|
| **RxNorm** | 205678 | Fluvoxamine 50 MG Oral Tablet [Luvox] |
| **RxNorm** | 205679 | Fluvoxamine 100 MG Oral Tablet [Luvox] |
| **RxNorm** | 335270 | nefazodone 25 MG |
| **RxNorm** | 763195 | {14 (nefazodone 100 MG Oral Tablet [Serzone]) / 14 (nefazodone 200 MG Oral Tablet [Serzone]) / 14 (nefazodone 50 MG Oral Tablet [Serzone]) } Pack [Serzone Starter Pack] |
| **RxNorm** | 335301 | Paroxetine 25 MG |
| **RxNorm** | 564737 | Trazodone 10 MG/ML [Molipaxin] |
| **RxNorm** | 565263 | Paroxetine 2 MG/ML [Seroxat] |
| **RxNorm** | 335300 | Paroxetine 12.5 MG |
| **RxNorm** | 825430 | Sertraline 0.2 MG/ML [Zoloft] |
| **RxNorm** | 825404 | Sertraline 0.2 MG/ML Oral Solution |
| **RxNorm** | 825403 | Sertraline 0.2 MG/ML |
| **RxNorm** | 1430124 | Paroxetine 7.5 MG [Brisdelle] |
| **RxNorm** | 1430120 | Paroxetine 7.5 MG |
| **RxNorm** | 406238 | Fluoxetine / olanzapine Oral Capsule [Symbyax] |
| **RxNorm** | 379510 | Fluoxetine Enteric Coated Capsule [Prozac Weekly] |
| **RxNorm** | 404733 | Fluoxetine 50 MG / olanzapine 6 MG Oral Capsule [Symbyax] |
| **RxNorm** | 404730 | Fluoxetine 25 MG / olanzapine 6 MG Oral Capsule [Symbyax] |
| **RxNorm** | 404732 | Fluoxetine 50 MG / olanzapine 12 MG Oral Capsule [Symbyax] |
| **RxNorm** | 404731 | Fluoxetine 25 MG / olanzapine 12 MG Oral Capsule [Symbyax] |
| **RxNorm** | 439544 | Paroxetine Extended Release Oral Capsule |
| **RxNorm** | 433852 | Paroxetine 25 MG 24 Hour Extended Release Tablet |
| **RxNorm** | 359852 | Paroxetine 37.5 MG 24 Hour Extended Release Tablet |
| **RxNorm** | 369962 | Paxil CR Extended Release Tablet |
| **RxNorm** | 352187 | Paroxetine 37.5 MG Extended Release Tablet [Paxil CR] |
| **RxNorm** | 379199 | Paroxetine 24 Hour Extended Release Tablet |
| **RxNorm** | 351867 | Paroxetine 12.5 MG Extended Release Tablet [Paxil CR] |
| **RxNorm** | 312939 | Sertraline 20 MG/ML Oral Solution |
| **RxNorm** | 359851 | Paroxetine 12.5 MG 24 Hour Extended Release Tablet |
| **RxNorm** | 331494 | Sertraline 20 MG/ML |
| **RxNorm** | 349055 | Sertraline 20 MG/ML Syrup |
| **RxNorm** | 386104 | Paroxetine Oral Solution [Seroxat] |
| **RxNorm** | 378265 | Sertraline Syrup |
| **RxNorm** | 284396 | Sertraline 20 MG/ML Oral Solution [Zoloft] |
| **RxNorm** | 369135 | Trazodone Oral Tablet [Desyrel Dividose] |
| **RxNorm** | 351868 | Paroxetine 25 MG Extended Release Tablet [Paxil CR] |
| **RxNorm** | 576615 | Fluoxetine 25 MG / olanzapine 12 MG [symbyax] |
| **RxNorm** | 576617 | Fluoxetine 50 MG / olanzapine 6 MG [symbyax] |

| | | |
|---|---|---|
| **RxNorm** | 576616 | Fluoxetine 50 MG / olanzapine 12 MG [symbyax] |
| **RxNorm** | 576614 | Fluoxetine 25 MG / olanzapine 6 MG [symbyax] |
| **RxNorm** | 575632 | Paroxetine 12.5 MG [Paxil CR] |
| **RxNorm** | 575918 | Paroxetine 37.5 MG [Paxil CR] |
| **RxNorm** | 575633 | Paroxetine 25 MG [Paxil CR] |
| **RxNorm** | 539512 | Sertraline Syrup [Zoloft] |
| **RxNorm** | 539511 | Sertraline 20 MG/ML [Zoloft] |
| **RxNorm** | 725079 | Fluoxetine / olanzapine Oral Capsule [Symbyax 3/25] |
| **RxNorm** | 700399 | Fluoxetine 4 MG/ML Syrup |
| **RxNorm** | 629007 | Fluoxetine 20 MG Oral Tablet [Prozac] |
| **RxNorm** | 725067 | Fluoxetine / olanzapine Oral Capsule [Symbyax 12/50] |
| **RxNorm** | 539513 | Sertraline 20 MG/ML Syrup [Zoloft] |
| **RxNorm** | 562161 | Paroxetine Oral Solution |
| **RxNorm** | 579897 | Fluoxetine Extended Release Oral Capsule |
| **RxNorm** | 579899 | Fluoxetine Extended Release Oral Capsule [Prozac Weekly] |
| **RxNorm** | 583057 | Fluoxetine Extended Release Oral Capsule [Prozac] |
| **RxNorm** | 700398 | Fluoxetine Syrup |
| **RxNorm** | 596854 | Fluoxetine 90 MG Extended Release Enteric Coated Capsule |
| **RxNorm** | 596853 | Fluoxetine Extended Release Enteric Coated Capsule |
| **RxNorm** | 721789 | Fluoxetine 25 MG / olanzapine 3 MG Oral Capsule [Symbyax] |
| **RxNorm** | 721788 | Fluoxetine 25 MG / olanzapine 3 MG [Symbyax] |
| **RxNorm** | 725075 | Fluoxetine / olanzapine Oral Capsule [Symbyax 12/25] |
| **RxNorm** | 725071 | Fluoxetine / olanzapine Oral Capsule [Symbyax 6/50] |
| **RxNorm** | 2532163 | PMDD fluoxetine 20 MG Oral Tablet |
| **RxNorm** | 2532159 | PMDD fluoxetine 10 MG Oral Tablet |
| **RxNorm** | 1182489 | Prozac Weekly Pill |
| **RxNorm** | 1185504 | Symbyax 6/25 Oral Product |
| **RxNorm** | 1185507 | Symbyax 6/50 Pill |
| **RxNorm** | 1180766 | Seroxat Oral Product |
| **RxNorm** | 1180765 | Seroxat Oral Liquid Product |
| **RxNorm** | 1185502 | Symbyax 3/25 Oral Product |
| **RxNorm** | 1182488 | Prozac Weekly Oral Product |
| **RxNorm** | 1185503 | Symbyax 3/25 Pill |
| **RxNorm** | 1185500 | Symbyax 12/50 Oral Product |
| **RxNorm** | 1185505 | Symbyax 6/25 Pill |
| **RxNorm** | 1185501 | Symbyax 12/50 Pill |
| **RxNorm** | 1185506 | Symbyax 6/50 Oral Product |
| **RxNorm** | 2591785 | citalopram 30 MG |
| **RxNorm** | 2591786 | citalopram 30 MG Oral Capsule |
